# Planning healthier cities by reframing the urban food landscape: measuring localised macronutrient exposure in Singapore

**DOI:** 10.1101/2025.04.10.25325565

**Authors:** Pei Ma, Shihui Jin, Yi Zhen Chew, Haolong Song, Borame Sue Lee Dickens

**Affiliations:** Saw Swee Hock School of Public Health, National University of Singapore and National University Health System, Singapore

**Keywords:** urban health, spatial epidemiology, built environment, macronutrient exposure, food environment, BMI

## Abstract

Rising overweight prevalence in Southeast Asian urban settings is contributing to substantial chronic disease burdens. The role of fast food outlet density or broader macronutrient-based exposure in these settings, as well as the applicability of international food environment measures, remains underexplored. Using Singapore as a case study representative of dense Southeast Asian cities such as Bangkok, Manila, Jakarta, Ho Chi Minh City, and Kuala Lumpur, we examine geospatial correlations between macronutrient exposure and overweightness rather than relying on categorical food outlet classifications. Data from 15,614 participants in Singapore’s Multi-ethnic Cohort were used to estimate macronutrient-based profiles of 14,764 geolocated food establishments and built environment covariates across 234 zones. Bayesian hierarchical models indicated that higher saturated fat exposure was associated with increased overweight risk (adjusted odds ratio [AOR] 1.11, 95% CrI 1.03–1.26), while greater exposure to green space was associated with reduced risk (AOR 0.65, 95% CrI 0.49–0.78). When we adjusted for multilevel spatial dependence, high saturated fat score remained significant. The association between overweight status and fast food outlet density was not statistically significant (AOR 1.00, 95% CrI 0.95–1.04). These findings suggest that spatial variation in nutritional exposure is associated with overweight risk beyond measures of fast food outlet density in Singapore, highlighting the need for more tailored and region specific urban planning approaches that consider the macronutrient landscape across similar cities in the wider region, potentially focusing on the availability of saturated fat in the local food environment.

## Introduction

The concurrent rise in global urbanisation and obesity prevalence represents a major challenge for public health and urban sustainability in the 21st century^1,2^. According to the World Health Organisation, obesity now affects over 1 billion people worldwide, with prevalence doubling between 1980 and 2022^3^. Whilst the causes of this rise are complex in etiology^4^, including non-modifiable risk factors such as age, sex and genetics^5^, the speed and scale of this epidemiological transition point to strong environmental drivers that extend beyond individual lifestyle choices^1^. Increasing attention has been directed toward obesogenic environments^6^, characterised by widespread availability of energy-dense, nutrient-poor ultra-processed foods, targeted marketing that promotes hedonic consumption^7^, and persistent barriers that limit access to healthier food options^8^. Importantly, exposure to obesogenic factors varies systematically across urban areas and is associated with spatial inequalities in health outcomes^9^. Addressing these differences requires approaches that extend beyond individual-level interventions and instead consider the spatial distribution of nutritional exposures within the urban environment^10,11^.

These structural dynamics are particularly evident in the rapidly urbanising regions of Southeast Asia, where accelerating commercial development has reshaped food retail landscapes with over 74,000 convenience stores emerging as of 2018^12^. Unlike traditional fresh markets, these modern outlets facilitate the increasing demand for ultra-processed products^13^. In Thailand, a substantial proportion of the population now regularly consume sugar-sweetened beverages (76.2%), packaged ready-to-eat foods (59.3%), and savoury snacks (48.3%)^14^. Projections for 2021–2026 also indicate a continuing 122% growth in expenditure on such products^14^. Similarly, Indonesia has recorded a large increase in the percentage contribution of fat towards caloric intake, from 11% to 23% between 1983 and 1999, corresponding with the region’s steepest rise in adult overweight prevalence between 1980 and 2016^15^. Singapore has observed similar trends, with average total dietary fat intake increasing from 94g in 2019 to 100g in 2022, 36% of which consists of saturated fat, exceeding the 30% recommendation^16^. National Nutrition Surveys further indicate that mean calorie intake has also shown an increase from 2360kcal in 2019 to 2410kcal in 2022, with 61% of individuals exceeding their daily recommended calorie intake, even after factoring for physical activity^16^. Collectively, these patterns illustrate an ongoing regional nutrition transition characterised by increasing fat consumption and declining carbohydrate intake, consistent with a shift away from traditional Asian dietary profiles and toward more energy-dense food environments.

Despite the well-established influence of demographic and socio-economic factors on health^17^, the independent role of the food environment remains difficult to quantify. Geospatial disparities, largely derived from Western contexts, rely on proxies such as the presence, proximity, or density of fast-food outlets, supermarkets, or “healthy” and “unhealthy” classified food retailers to infer caloric exposure. Yet, within Southeast Asia, these metrics have produced inconsistent associations with obesity^18^, possibly due to different cultural contexts^19^. For example, Kway Chap, a popular local dish in Singapore, contains approximately 650 kcal per portion and 12 grams of saturated fat, both of which are higher than many fast food burgers^20,21^. Considerations on contextualised Southeast Asian food environment metrics are thus required to assess their potential role in rising obesity and overweight prevalence in their populations. With limited representative data on nutritional intake and availability across the region, we devise an alternative approach to create a food environment index to assess the potential impact of the local food environment on an individual’s health.

Here we use Singapore as a case study of major Southeast Asian cities, which are characterised by very high population densities within relatively small urban footprints, compact urban form, strong land-use mixing, and a predominance of vertical development. Comparable urban characteristics can be observed in other large Southeast Asian cities such as Bangkok, Manila, Jakarta, Ho Chi Minh City, and Kuala Lumpur^22^. This study utilises anthropometric data from the Multi-ethnic Cohort^23^ (n□=□15,614) and 14,764 geolocated food establishments and built environment covariates across 234 zones. We aim to: (1) characterise the food environment beyond outlet typologies by quantifying macronutrient-based availability using systematic web-based data extraction from publicly available online menus of food establishments in Singapore, identified through a national postcode-linked registry and supplemented with estimated nutritional content for local food stalls based on standardised meals and previously measured nutritional data; (2) examine correlations between this macronutrient-based metric with conventional fast-food outlet density measures, and (3) estimate geospatial inequalities in fast-food outlet density and macronutrient-based availability alongside their associations with area-level prevalence of overweight and obesity.

## Methods

### Overview

This study adopted a spatial structure and proceeded in three stages. First, multi-scale geospatial data were integrated to quantify built and food environment characteristics at the spatial-unit level across Singapore and mapped to resident-level exposures. Second, spatially constrained clustering was applied to the built environment, food environment, and socio-economic indicators to identify neighbourhood typologies across Singapore. Third, multi-level logistic regression models were used to evaluate associations between individual overweight status and environmental exposures, while accounting for socio-demographic factors and spatial heterogeneity. A description of all co-variates explored is provided (Table 1).

**Table 1.** Candidate predictors for univariable regression models.

| Variable | Definition |
| --- | --- |
| **Socio-demographics** |  |
| Age (group) | Categorical, including 21–24, 25–34, 35–44, 45–54, 55–64 and 65+; Age defined as years from birth to the time of the MEC2 study |
| Gender | Categorical, including male and female |
| Ethnicity | Categorical, including Chinese, Malays, and Indians |
| Marital Status | Categorical, including single, married, divorced/separated/widowed |
| High education | Whether they have received university education |
| Long education | Whether they have received at least 11 years of education (equivalent to completing A-level education in the Singaporean context) |
| Work status | Categorical, including working population, student, homemaker, retired, and unemployed status |
| Smoking status | Categorical, including never smoked, ex-smoker, current occasional smoker, and current daily smoker |
| Hypertension | Presence of hypertension diagnosis by a western-trained medicine doctor |
| Total physical activity level | Categorical, including 0 to <50, 50 to <100, 100 to <150, 150+ (met-hr/week) |
| **Food environment** |  |
| Calorie index | Average calorie content of meals within a spatial unit, transformed into a quantile-based, integer-valued index ranging from 0 (lowest) to 10 (highest) |
| Carbohydrate index | Carbohydrate content of meals within a spatial unit, transformed into a quantile-based, integer-valued index ranging from 0 (lowest) to 10 (highest) |
| Fat index | Fat content of meals within a spatial unit, transformed into a quantile-based, integer-valued index ranging from 0 (lowest) to 10 (highest) |
| Saturated fat index | Saturated fat content of meals within a spatial unit, transformed into a quantile-based, integer-valued index ranging from 0 (lowest) to 10 (highest) |
| Protein index | Protein content of meals within a spatial unit, transformed into a quantile-based, integer-valued index ranging from 0 (lowest) to 10 (highest) |
| High saturated fat score | Proportion of food retailers where the saturated fat intake exceeded 10% of total calorie intake per standard meal provided within a spatial unit |
| Fast food outlet density | Number per square meter of fast food outlets within a spatial unit |
| **Built environment** |  |
| Density of intersections | Density of intersections within a spatial unit (intersections/m<sup>2</sup>) |
| Density of bus stops | Density of bus stops within a spatial unit |
| Density of MRT stations | Density of MRT stations within a spatial unit |
| Density of taxi stands | Density of taxi stands within a spatial unit |
| Walkway accessibility | Percentage of walkways accessible within a spatial unit |
| MRT<sup>#</sup> accessibility | Percentage of MRT lines accessible within a spatial unit |
| Highway accessibility | Percentage of highways accessible within the spatial unit |
| Proportion of commercial areas | Percentage of land within the spatial unit designated for commercial use |
| Proportion of residential areas | Percentage of land within the spatial unit designated for residential use |
| Abundance of green space | Categorical, with low and high levels in which green space comprises 0 to <19% and 19%+ of the spatial unit |

<sup>#</sup> MRT: The Mass Rapid Transit System, the major mode of railway transportation in Singapore.

### Food establishment data

We adapted the INFORMAS^24^ framework to collate near-complete data on 14,764 food establishments across 2,655 postal codes in Singapore, covering 26 categories of commonly observed food establishments including restaurants, cafes, hawker centres, convenience stores, and vending machines, to characterise neighbourhood food availability. Lists of food establishments were retrieved through QuickOSM queries from the OpenStreetMap platform^25^ and subsequently validated against Google Maps^26^ and the Singapore OneMap^27^ database to ensure positional and categorical accuracy. Attributes for each establishment included outlet type, brand, name, address, latitude, longitude, and a standardised food set profile. Data cleaning procedures involved removing duplicates, correcting misclassifications, and aligning all geographic coordinates with the national SVY21 projection.

### Food set profile data

Publicly available menu information reflecting actual food offerings was extracted from food establishments’ official websites and Google Maps profiles. These data were used to identify typical food compositions for each outlet and to construct representative food set profiles across different food categories. For example, the standard food set profile for a Hainanese chicken rice restaurant consists of six components: chicken fillet, chicken rice, cucumber slices, spring onion, chicken soup, and chilli sauce. Outlets of this type were assumed to provide meals consistent with this standardised composition, representing the meal typically consumed by individuals visiting such establishments. To ensure consistency and comparability, all outlets of the same type were assigned a single representative food set profile across Singapore. In total, 525 distinct food set profiles were compiled, each corresponding to a unique food outlet category. The consistency of this classification was subsequently validated through cross-referencing multiple menu sources and collector review.

### Nutrient composition data

Nutritional information was retrieved for each food composition from three authoritative sources: the USDA^28^ Nutritive Value Booklet, the USDA FoodData Central online database^28^, and Singapore’s Health Promotion Board (HPB) Energy and Nutrient Composition of Food Database^29^. When macronutrient-based data for specific local items were unavailable in these primary references, supplementary verification was performed using reputable online nutrition databases to ensure completeness and consistency. For each component, standardised macronutrient-based values were scaled by the ratio of actual to standard portion weight, to derive proportional macronutrient-based contributions. We focused on five key macronutrient-based factors: calories, total fat, saturated fat, protein, and carbohydrates. For instance, a typical 330 g serving of Hainanese chicken rice was estimated to provide approximately 524.72 kcal, 21.78 g protein, 19.75 g total fat, 7.55 g saturated fat, and 64.47 g carbohydrates.

### Nutrient-related indices

By integrating data on food establishments, food set profiles, and macronutrient-based compositions, we created the first high-resolution dataset of macronutrient-based spatial distribution across Singapore, providing a measure of residents’ local exposure to nutrition.

The city-state was divided into 954 spatial units representing discrete neighbourhood areas delineated by major roads or commercial land use. For each spatial unit *i*, the value of macronutrient-based variable *n* (calories, total fat, saturated fat, protein, or carbohydrates) was aggregated by summing the corresponding values across all food establishments and then dividing the summed value by the number of postal codes within the unit, i.e.,

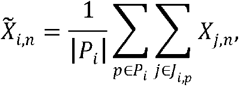

where *χ*_*j,n*_, denotes the value of factor *n* for establishment *j*; *J*_*i,p*_, represents the set of establishments located in postcode *p* within spatial unit *i*; and | *P*_*i*_ | is the total number of postal codes in that unit. Across all spatial units, zero 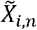, values were assigned a value of 0, and the remaining non-zero values were normalised and grouped into ten quantile bins to derive macronutrient-based indices:

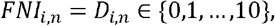

where higher values reflected greater localised exposure intensity.

The high saturated fat score was also computed for each spatial unit, defined as the proportion of food establishments within spatial unit *i* for which saturated fat accounted for at least 11% of total caloric content, consistent with World Health Organization guidelines. This score was designed to capture the structural composition of the local food environment by distinguishing high saturated fat options from foods with higher overall macronutrient-based availability alone.

These spatially aggregated variables were subsequently linked to individual cohort participants based on their residential addresses.

### Built environment data

The built environment framework was developed from an urban accessibility perspective, linking the spatial structures of transport systems and land use. Urban form was characterised by points (transport nodes), lines (transport flows), and polygons (functional zones) to capture interactions between the transport system and built-up land-use, reflecting spatial accessibility within Singapore’s urban environment.

Point and line features were obtained from OpenStreetMap extracts via the GEOFABRIK repository^30^ updated in March 2024, and polygonal land-use data were sourced from the Urban Redevelopment Authority’s Master Plan 2019 Land Use dataset^31^. Three quantitative attributes were derived: (i) point density, defined as the number of transport nodes per square kilometre including intersections, bus stops, MRT stations and taxi stands; (ii) line proportion, representing the share of total network length by class including walkways, MRT lines and highways; and (iii) polygonal coverage, quantified by the proportion of commercial, residential and green land-use area.

Built environment variables were derived using the same spatial partitioning applied in calculating nutrient-related factors, enabling their direct mapping to individual cohort participants. For each spatial unit *i*, indicators were calculated as:

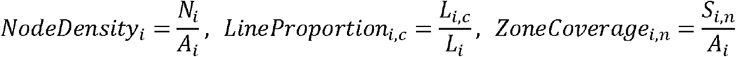

where *N*_*i*_ is the number of nodes, *L*_*i,c*_, is the length of transport type *c, L*_*i*_ is the total road length, *S*_*i,n*_ is the area of land-use type *n*, and *A*_*i*_ is the total area.

### Socio-demographic and anthropometry data

De-identified individual-level socio-demographic, anthropometric, and geospatial data were obtained from the Multi-Ethnic Cohort Phase 2 (MEC2)^23^ follow-up study, conducted between 2017 and 2021 as part of the Singapore Population Health Studies by the National University of Singapore. The MEC2 cohort provides population-based data representative of Singapore’s major ethnic groups and adult age range. A total of 19,700 participants were recruited through standardised home visits across all planning regions in Singapore, and written informed consent was obtained before participation.

Socio-demographic variables included age, gender, ethnicity, education level, years of education, marital status, employment status, average monthly earnings, housing type, smoking status, hypertension status, and total physical activity level (Table S1). Education level and income were self-reported, while physical activity level was assessed using standardised questionnaires. Anthropometric measurements, including height, weight, and blood pressure, were taken by trained field staff using calibrated equipment following standardised protocols. Body mass index (BMI) was calculated as weight in kilograms divided by height in meters squared (kg/m^2^). Overweight status was defined according to the Asian BMI threshold of 23 kg/m^2^ or higher, encompassing both overweight and obesity. Hypertension was defined as systolic blood pressure of 140 mmHg or higher or diastolic blood pressure of 90 mmHg or higher.

Each participant’s residential address was geocoded and mapped to one of the 954 predefined spatial units in Singapore using the national SVY21 coordinate system. A total of 4,086 participants were excluded due to missing data, non-major ethnicity classification, or residence in spatial units with fewer than three respondents. The final analytic sample consisted of 15,614 participants, distributed across 234 spatial zones.

### Spatially constrained clustering of spatial zones

Spatially constrained hierarchical clustering was applied to the 954 spatial zones in Singapore to capture spatial disparities in macronutrient exposure, built environment, and socio-economic conditions. Ten spatial-unit-specific indicators were included: the five nutrient-related indices, high saturated fat score, outlet diversity (number of distinct retailer types within individual spatial units), green space proportion, housing price (average price of public housing within individual spatial units), and population density. All variables were standardised to have a mean of 0 and a standard deviation of 1, and Euclidean distances between spatial units were then calculated based on these features. Together with geographic distances between unit centroids, the distance matrix was utilized to perform Ward clustering, implemented through the ClustGeo^32^ R package. Hyperparameters, including the spatial constraint weight *α* (= 0.45) and cluster count *K* (= 5), were selected using grid search to balance within-cluster attribute homogeneity and spatial coherence (Figure S6–S7).

### Logistic regression analyses

We constructed a series of multi-level logistic regression models to investigate associations between individual overweight status and exposure to built and food environments, adjusting for socio-demographic characteristics and spatial heterogeneities. Overweight status was modelled as a binary outcome, and all continuous predictors were standardised to a zero mean and unit variance before analysis. Parameters were estimated under a Bayesian framework using the brms^33^ R package with Hamiltonian Monte Carlo sampling. Weakly informative priors of N(0, 2.5) were assigned to all fixed-effect coefficients. Four chains were run in parallel, each with 2,000 warm-up iterations followed by 4,000 sampling iterations. Model convergence was assessed using visual inspection of trace plots, the Gelman-Rubin convergence diagnostic (R-hat <1.01), and effective sample size (>1,000). Parameter estimates were reported as posterior medians and 95% credible intervals, summarised from the resulting 16,000 posterior draws.

### Univariable models

We first employed univariable regression analyses to select candidate predictors for multivariable regression models. For each predictor *χ*, its relationship with the risk of being overweight *p* was modelled as

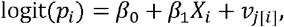

where *v*_*j*[*i*]_ is the random intercept for the spatial unit *j* to which the individual *i* belongs, with 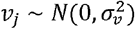. Predictors with statistically significant odds ratio estimates (i.e., 95% CrIs not containing 1) were retained for subsequent multivariable analyses (Table 2).

**Table 2.** Posterior median odds ratios and 95% credible intervals (CrIs) for candidate predictors estimated from univariable logistic regression models.

|  | Predictor | Odds ratio | 95% CrI |
| --- | --- | --- | --- |
|  | **Socio-demographics** |  |  |
| Gender | Male (baseline) | 1 |  |
|  | Female | 0.62 | [0.58,0.66] |
| Age | 21–24 | 0.46 | [0.36,0.60] |
|  | 25–34 | 0.71 | [0.62,0.80] |
|  | 35–44 | 1.16 | [1.03,1.30] |
|  | 45–54 | 1.29 | [1.16,1.43] |
|  | 55–64 | 1.19 | [1.06,1.32] |
|  | 65+ (baseline) | 1 |  |
| Ethnicity | Chinese (baseline) | 1 |  |
|  | Malays | 2.85 | [2.57,3.17] |
|  | Indians | 3.21 | [2.91,3.55] |
| High education | No (baseline) | 1 |  |
|  | Yes | 0.78 | [0.73,0.83] |
| Long education | No(baseline) | 1 |  |
|  | Yes | 0.90 | [0.87,0.93] |
| Marital Status | Single (baseline) | 1 |  |
|  | Married | 1.85 | [1.71,2.01] |
|  | Divorced/ Separated/Widowed | 1.80 | [1.58,2.06] |
| Work status | Working (baseline) | 1 |  |
|  | Student | 0.37 | [0.31,0.46] |
|  | Homemaker | 1.07 | [0.97,1.18] |
|  | Retired | 0.84 | [0.72,0.96] |
|  | Unemployed | 0.82 | [0.70,0.96] |
| Smoking Status | Never smoked (baseline) | 1 |  |
|  | Ex-smoker | 1.72 | [1.51,1.96] |
|  | Current occasional smoker | 1.35 | [1.11,1.66] |
|  | Current daily smoker | 1.17 | [1.06,1.29] |
| Hypertension | No(baseline) | 1 |  |
|  | Yes | 2.60 | [2.36,2.87] |
|  | Total physical activity level (per 10 met-hr/wk) | 1.17 | [1.13,1.21] |
|  | **Food environment** |  |  |
|  | Calorie index | 1.08 | [1.02,1.14] |
|  | Carbohydrate index | 1.07 | [1.01,1.13] |
|  | Fat index | 1.07 | [1.02,1.13] |
|  | Saturated fat index | 1.07 | [1.01,1.13] |
|  | Protein index | 1.06 | [1.01,1.12] |
|  | High saturated fat score | 1.12 | [1.05,1.20] |
|  | Fast food outlet density | 1.00 | [0.95, 1.04] |
| **Built environment** |  |  |  |
|  | Density of intersections | 1.00 | [0.96,1.05] |
|  | Density of bus stops | 1.04 | [0.99,1.09] |
|  | Density of MRT stations | 0.98 | [0.94,1.03] |
|  | Density of taxi stands | 0.94 | [0.90,0.98] |
|  | Walkway accessibility | 1.01 | [0.96,1.06] |
|  | MRT<sup>#</sup> accessibility | 0.98 | [0.94,1.03] |
|  | Highway accessibility | 1.02 | [0.96,1.07] |
|  | Proportion of commercial areas | 0.97 | [0.93,1.02] |
|  | Proportion of residential areas | 1.00 | [0.95,1.06] |
| Abundance of green space | 0 to <19% (baseline) | 1 |  |
|  | 19%+ | 0.59 | [0.48,0.70] |

### Multivariable models

In the baseline model, six predictors were assessed, including gender, age group, ethnicity, highest education, work status, hypertension status, and green space exposure. We additionally assessed 6 model variants, each with either one of the nutrient-related indices or high saturated fat score as the additional predictor, allowing for evaluation of their adjusted associations with overweight risk.

As in the univariable models, a spatial-zone-specific random intercept was included to account for unobserved spatial heterogeneity. Each multivariable model took the form:

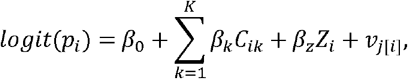

where *C*_*ik*_ denotes the *k*-th baseline covariate value for the individual *i* and *Z*_*i*_ represents the nutrient-related exposure of interest for that individual.

### Multivariable models with cluster-level random effects

To assess whether the spatially constrained clustering adequately captured spatial heterogeneities, we extended the previous multivariate model by incorporating a cluster-level random intercept and spatial autocorrelation:

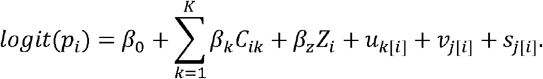

In this, *u*_*k*[*i*]_ denotes the random intercept for the cluster *k* to which individual *i* belongs, with 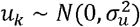. Structured spatial dependence was captured using *s*_*k*[*i*]_, which was assigned an intrinsic conditional autoregressive prior *s*_*j*_ | 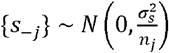, with *n*_*j*_ being the number of neighbours. A constraint ∑_*j*_*s*_*j*_ =0 was additionally imposed to ensure model identifiability.

## Results

### Built and food environment covariates

Data on 14,764 food establishments were collected across 2,655 postal codes nested within 945 spatial units in Singapore (Figure S1). A total of 525 standardised food set profiles, representing typical meals served at each establishment, were compiled from published menus, with five nutrient-related factors, including calories, carbohydrates, total fat, saturated fat, and protein, calculated based on the corresponding food components. Each spatial unit contained up to 172 distinct food set profiles with regional disparities observed for dietary diversity, food establishment numbers and fast-food outlet density (Figure S2-S4). The aggregated nutritional profiles also varied across spatial units (Figure 1), with substantial variation observed in macronutrient profiles (Figure 1, Figure S5), which did not align with population density or built environment features (Figure 2).

**Figure 1.**
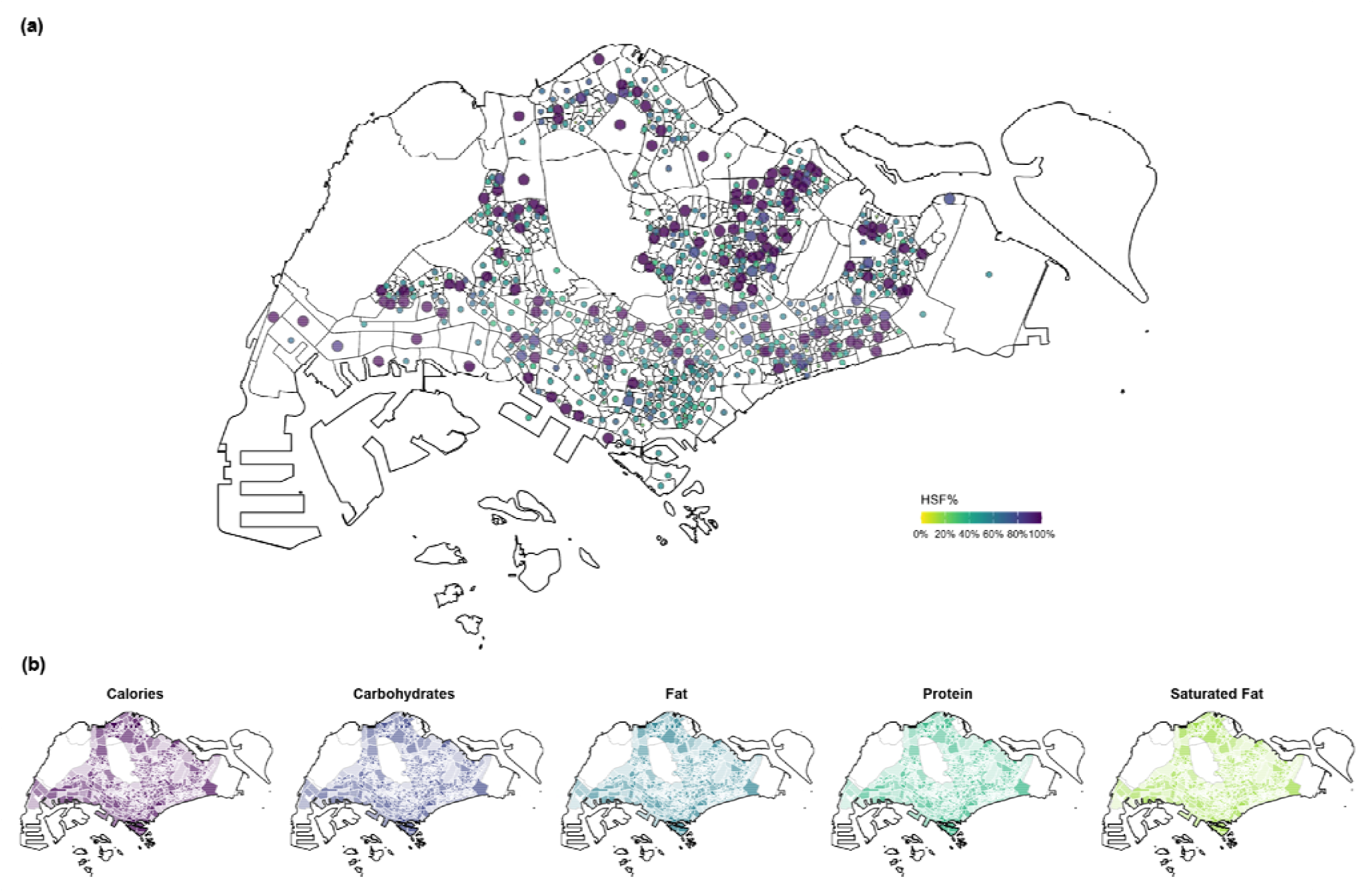
Spatial distribution of nutrient exposure. Subfigure (a) shows the high saturated fat score (continuous, 0–100%, representing the percentage of outlets serving meals with at least 11% saturated fat content) across the 954 spatial units in Singapore, with higher values represented by larger circles and darker colours. Subfigure (b) presents five nutrient-related factors at the spatial-unit level, each defined as a quantile-based, integer-valued index ranging from 0 to 10.

**Figure 2.**
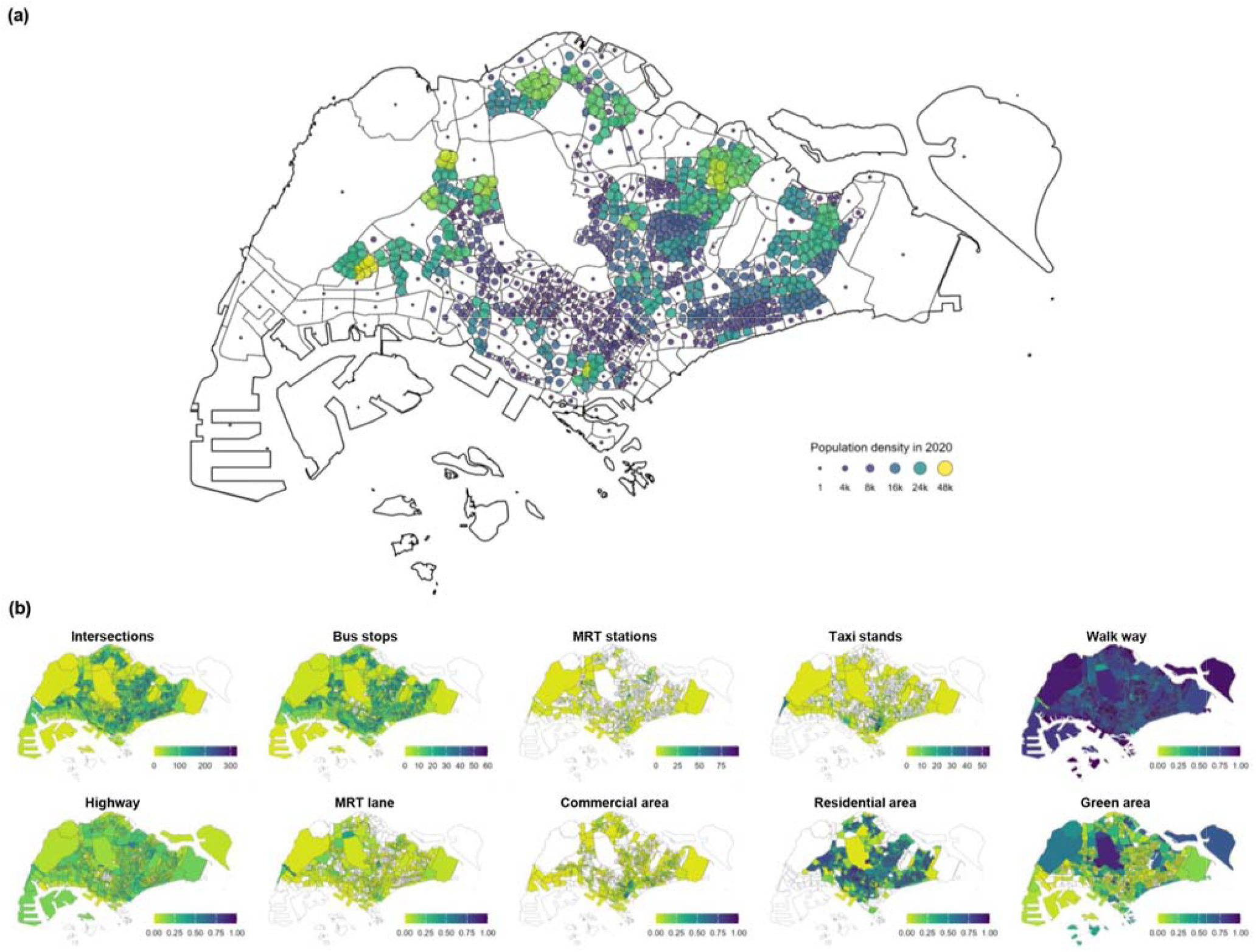
Spatial patterns of (a) population density and (b) built environment indicators across Singapore. In subfigure (a), larger circles and lighter colours indicate higher population density of the relevant spatial unit. In subfigure (b), darker colours denote higher densities of the corresponding built environment factors.

### Spatial clustering of built, food, and socio-economic factors

Spatially constrained clustering was applied to classify the 954 spatial units into five neighbourhood clusters (A–E), corresponding broadly to southern, western, eastern, northern, and central Singapore. These clusters exhibited distinct socio-demographic, built, and food environment characteristics (Figure 3). Spatial units in Cluster E generally exhibited lower values across the five nutrient-related indices. However, their high saturated fat scores, defined as the percentage of food outlets within a spatial unit where saturated fat contributed at least 11% of the total caloric content, were comparable to those in other clusters. Meanwhile, outlet diversity in these units was relatively low, suggesting a more limited food supply. Cluster D was characterised by higher population density and lower housing prices, alongside greater green space coverage. In contrast, spatial units in Cluster A, many of which were designated for commercial land use, were characterised by higher outlet diversity and housing prices but substantially lower population density. Higher high saturated fat scores were observed among spatial units in Cluster C, while those in Cluster B showed no pronounced deviations from citywide averages across the evaluated factors (Figure 3).

**Figure 3.**
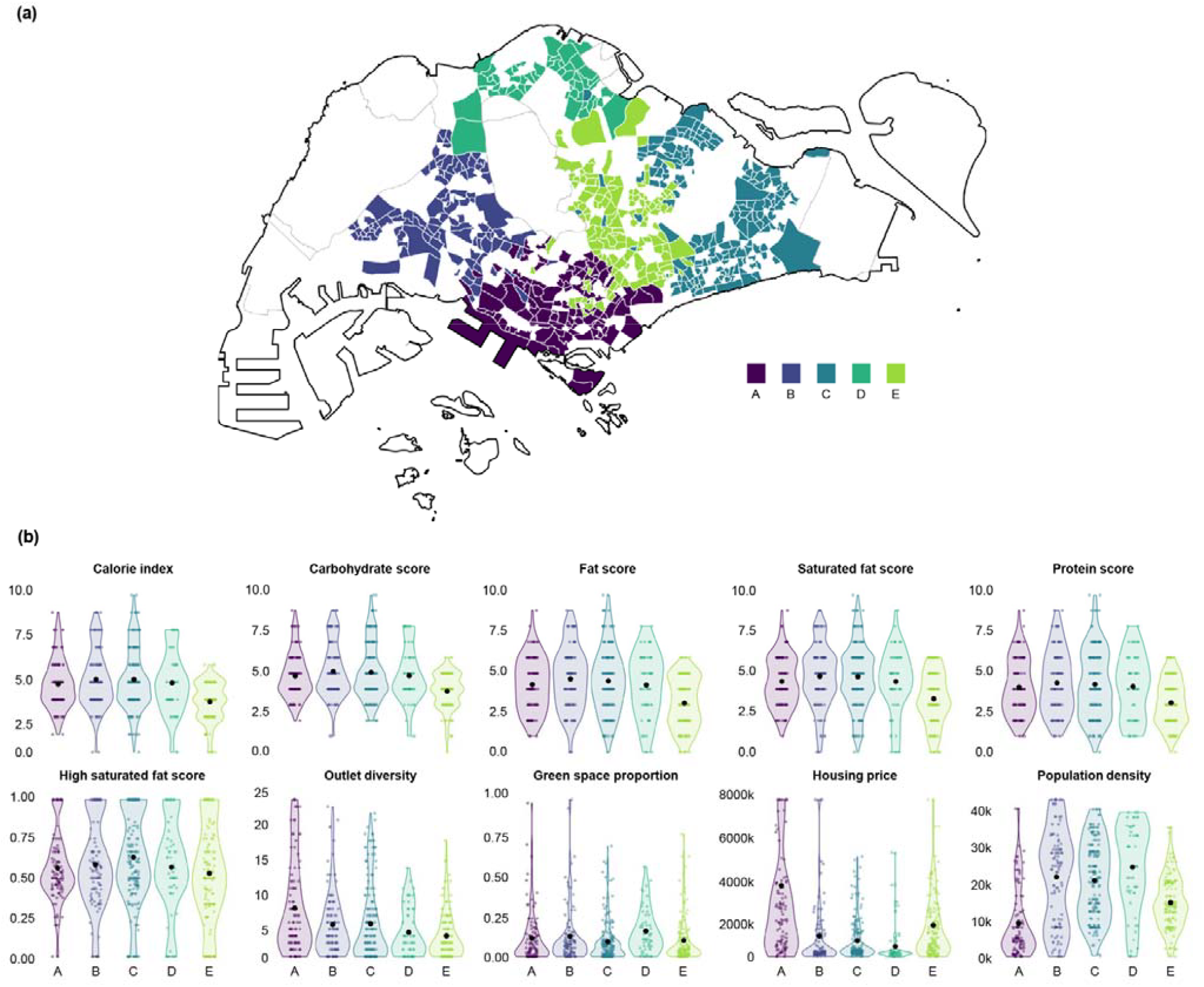
Clusters identified using spatially constrained clustering. (a) Spatial distribution of the five clusters across Singapore, showing the spatial units assigned to each cluster. (b) Distributions of spatial-unit–level values for the ten indicators used in clustering, stratified by cluster. Clusters are distinguished by colour. The black points represent the cluster-specific mean values for individual indicators.

**Figure 4.**
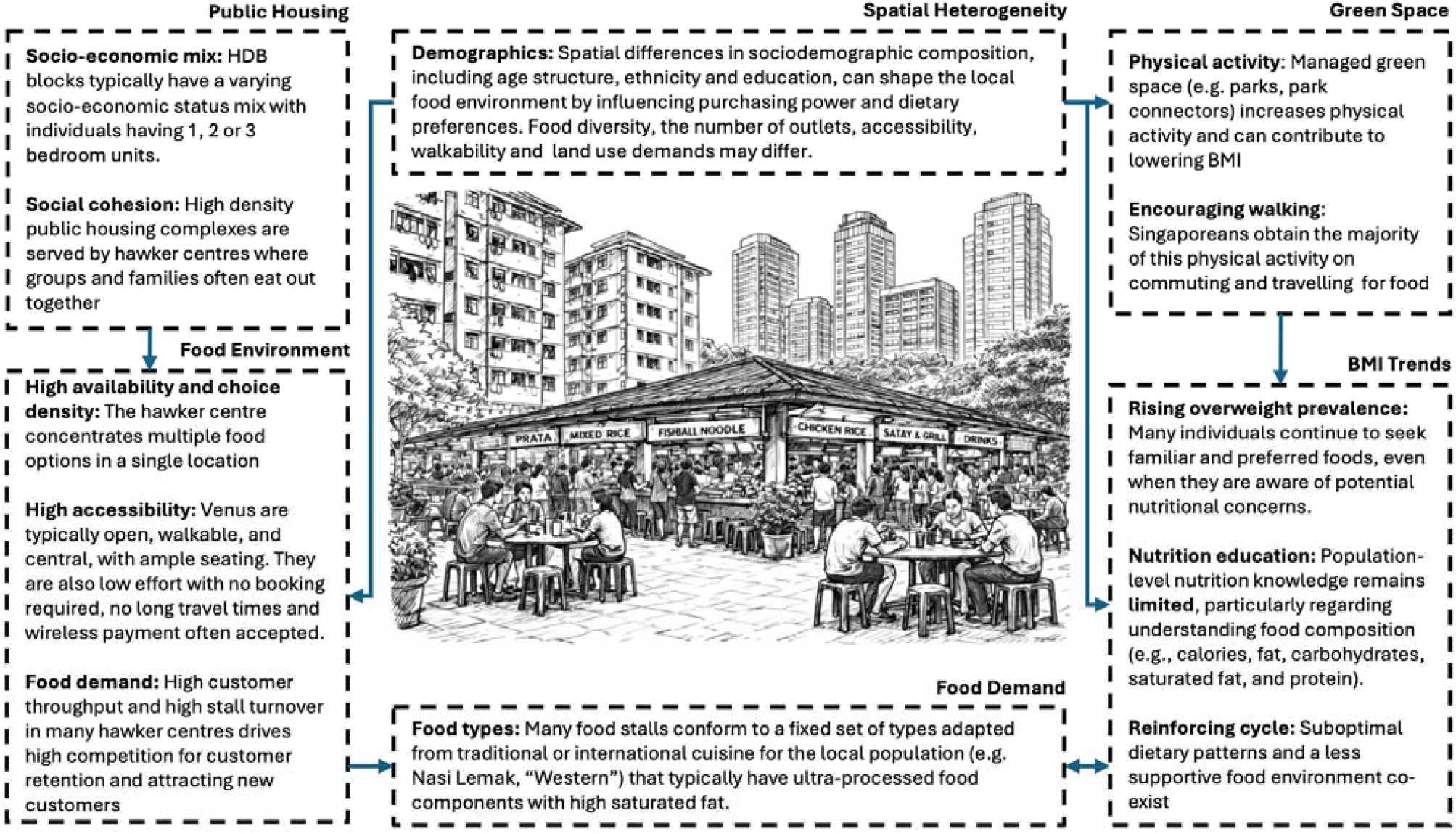
Conceptual framework linking urban context, food environment, and BMI patterns in Singapore. The diagram illustrates how the public housing system and spatial heterogeneity shape local food demand, food availability, and physical activity opportunities. These upstream structural factors interact with dietary choices, nutritional knowledge, and commuting-related activity patterns, jointly influencing population-level BMI trends in the Singapore urban context.

### Socio-demographic and anthropometric characteristics of participants

A total of 15,614 individuals were included in this study (Table S1), of whom 9,548 (61.2%) were classified as overweight (BMI ≥23 kg/m^2^) and 6,066 (38.8%) as non-overweight. The age distribution was centred around middle adulthood, with the largest proportion aged 45–54 years (27.1%), followed by 55–64 years (22.4%) and 35–44 years (21.0%). Younger age groups were less represented, with 13.8% aged 25–34 years and 1.7% aged 21–24 year

Females comprised 54.2% of the sample, although overweight prevalence was higher among males (50.3% of overweight individuals vs 38.7% of non-overweight; p < 0.001). The majority of participants were Chinese (66.8%), followed by Indians (18.1%) and Malays (15.1%), broadly reflecting Singapore’s ethnic composition. Overweight prevalence differed significantly by ethnicity, with Indians (23.2%) and Malays (18.9%) representing larger proportions of the overweight group relative to Chinese participants (57.9%; p < 0.001).

Most participants were married (71.7%) and working (73.1%), indicating a predominantly working-age population. Socioeconomic indicators showed that 27.9% reported monthly household incomes between SGD 2,000 and <4,000, while 25.5% reported incomes below SGD 2,000. In terms of education, 22.9% had attained university-level education, while 77.1% had A-level qualifications or lower. Housing type distribution was largely concentrated in HDB flats, particularly 4-room flats (38.6%), though housing type was not significantly associated with overweight status (p = 0.28).

Health and behavioural characteristics also differed between groups. Hypertension was reported by 17.1% of participants, with a substantially higher prevalence among overweight individuals (21.7% vs 9.8%; p < 0.001). Smoking status varied significantly across weight categories, with overweight individuals more likely to be ex-smokers (9.2%) or current smokers compared with non-overweight participants (p < 0.001). Physical activity levels also differed, with overweight participants more frequently reporting ≥150 MET-hours per week (26.4%) compared with 21.8% among non-overweight individuals, although lower activity categories remained common across both groups (p < 0.001).

Significant differences between the overweight and non-overweight groups were observed for all the socio-demographic factors except housing type (Table 2). Characteristics associated with a higher overweight risk included middle age, male gender, ethnicities other than Chinese, lower income and lower education levels. A strong association was also observed between overweight status and individual physical health indicators. Individuals with a history of smoking, regardless of current smoking status, as well as those with hypertension or low physical activity levels, were more susceptible to becoming overweight.

### Built and food environment correlates of overweight status

The univariable regression analyses showed no evidence that individual overweight status was associated with fast food outlets density (odds ratio per additional fast-food outlet per square meter: 1.00, 95% credible interval [CrI]: 0.95– 1.04), after adjusting for spatial heterogeneity (Table 3). In contrast, statistically significant associations were estimated for all five nutrient-related indices and the high–saturated-fat score, which were therefore carried forward into multivariable regression analyses (Model 1–6).

**Table 3.** Estimated adjusted odds ratios (posterior medians and 95% CrIs) for the food and built environment predictors in Model 1–6. These six models respectively incorporated the calorie index, carbohydrate index, fat index, protein index, saturated fat index, and high saturated fat score as predictors in addition to socio-demographic factors and abundance of green space. Full model outputs (Model 1–Model 6) are available in Table S2.

| Predictor | Model 1 | Model 2 | Model 3 | Model 4 | Model 5 | Model 6 |
| --- | --- | --- | --- | --- | --- | --- |
| **High green space coverage** | 0.71<br>[0.60,0.81] | 0.71<br>[0.60,0.82] | 0.71<br>[0.60,0.81] | 0.71<br>[0.60,0.82] | 0.71<br>[0.60,0.82] | 0.65<br>[0.49,0.78] |
| **Calorie index** | 1.06<br>[1.01,1.12] |  |  |  |  |  |
| **Carbohydrate index** |  | 1.05<br>[0.99,1.11] |  |  |  |  |
| **Fat index** |  |  | 1.05<br>[1.00,1.12] |  |  |  |
| **Protein index** |  |  |  | 1.05<br>[0.99,1.11] |  |  |
| **Saturated fat index** |  |  |  |  | 1.05<br>[1.00,1.11] |  |
| **High saturated fat score** |  |  |  |  |  | 1.11<br>[1.03,1.26] |

Among the six retained predictors, the high saturated fat score demonstrated the strongest association with overweight status. Each one percentage point’s increase in this measure was inferred to be associated with an 11% (95% CrI: 3%–26%) increase in the odds of being overweight, holding the socio-demographic and built environment factors in the multivariable models constant. The relationship between overweight status and the indices for calories, fat, and saturated fat was also statistically significant, where a one-unit increase in these three indices was associated with a 6% (95% CrI: 1%–12%), 5% (95% CrI: 0%–12%), and 5% (95% CrI: 0%–11%) increase in overweight odds, respectively (Table 3).

The association between overweight status and green space availability remained consistent across Models 1–5. On average, individuals residing in spatial units with green space coverage exceeding 19% were estimated to be 29% (95% CrI: 18%–40%) less likely to be overweight after adjustment for socio-demographics and food-environment-related factors. This association increased to a 35% (95% CrI: 22%–51%) reduction in overweight odds when the high saturated fat score replaced the nutrient-related indices in the adjustment set, as shown in Model 6 (Table 3).

After adjusting for multilevel spatial dependence through clustered random effects and spatial autocorrelation, the association of overweight status with the five nutrient-related indices became statistically insignificant. In contrast, the association with high saturated fat score remained significant, with each percentage point’s increase in this measure linked with a 55% (95% CrI: 29%–149%) increase in the odds of being overweight (Table S3).

## Discussion

We examined associations between geospatial macronutrient profiles and body mass index across a cohort of 15,614 individuals, integrating high-resolution mapping data on 14,764 geolocated food establishments in Singapore. Analyses accounted for heterogeneity by age, ethnicity, sex, education status, high blood pressure status, and work status, as well as features of the built environment. Higher BMI and overweight risk were more common among middle-aged participants, individuals of Indian and Malay ethnicities, males, and those with lower educational attainment or non-working status. Lower local green space density was also associated with higher overweight risk, consistent with findings reported elsewhere^34^. In contrast, conventional fast food outlet density was not associated with overweight status; however, higher estimated macronutrient availability, including total fat, saturated fat, and carbohydrates, was associated with overweight prevalence, as was higher estimated total energy availability across the 234 spatial units in which cohort participants resided.

Globally, increasing portion sizes and rising meal energy content have been documented across food establishments, irrespective of outlet classification, and these trends have been linked to increasing overweight prevalence in multiple populations^35^. In a United Kingdom study of 27 restaurant chains and six fast food chains comprising 13 396 meals, the mean energy content was 977 kcal per meal, excluding drinks, snacks, and desserts^35^. In a separate study of 223 meals sampled from 111 food outlets across five countries, including Brazil, India, Finland, Ghana, and China, 94% of full-service meals and 72% of fast food meals contained at least 600 kcal, excluding desserts, snacks, and drinks^36^. These findings suggest that focusing exclusively on fast food establishments may underestimate the contribution of the broader food environment to excess energy intake. Consistent with this, the total energy content of fast food meals was significantly lower than meals from full-service restaurants in Brazil (34%; P=0.03), China (46%; P<0.001), and the United States (29%; P<0.001)^36^.

In Singapore, a macronutrient-based analysis of 45 commonly consumed meals across three major ethnic groups found that, if consumed three times daily, all meals exceeded recommended guidelines for total energy intake^20^. For example, a typical serving of laksa, a widely consumed coconut curry noodle dish, contains approximately 40% of the recommended daily energy intake, 40% of the recommended daily fat intake, and 30% of the recommended daily carbohydrate intake. As a result, maintaining energy intake within World Health Organisation recommended thresholds may be challenging, even when consuming foods that are not traditionally classified as fast food or unhealthy. Many commonly available meals resemble home-style dishes but remain energy dense, implying that interventions focused solely on conventional fast food outlets may have limited impact. However, implementing broad-based regulations to reduce energy density across the full range of food establishments would likely be costly and manpower intensive to design, implement, and enforce.

The effects of increasing meal energy density are further compounded by the high frequency of eating outside the home in many urban Southeast Asian populations. In these settings, commercial fast food outlets may be perceived as luxury food, while local foods or full-service restaurants are often viewed as healthier alternatives^37^. In Singapore, 77.3% of individuals report consuming at least one meal outside the home daily, typically at affordable establishments such as food courts, locally referred to as hawker centres. These outlets are convenient and low-cost, and do not necessarily rely on mass production for commercial resale. However, intense competition between vendors may incentivise the use of ingredients that enhance palatability, including higher saturated fat content and overall energy density^38^.

Higher saturated fat consumption, commonly derived from meat and dairy products, has been hypothesised to contribute to weight gain and adverse metabolic risk profiles^39^, mainly due to the relative energy density of fat at 9kcal/g compared to carbohydrate at 4kcal/g. Nevertheless, evidence on the long term impact of dietary macronutrient composition on weight gain remains mixed. Randomised trials evaluating low-fat and high fat dietary patterns have often faced challenges related to adherence and changes in overall diet quality, contributing to inconsistent conclusions regarding the role of dietary fat and carbohydrate composition in obesity risk^40–42^. In a United States cohort study of 41,518 women followed over eight years, total fat intake showed only a weak positive association with weight gain, while saturated fat intake was more strongly associated, with each percentage increase in caloric contribution corresponding to a 0.34 kg increase in weight^43^. In contrast, in a separate study of 136,432 participants with four-year changes assessed, increases of 100 g per day in starch, added sugar, and total carbohydrate intake were associated with weight gains of 1.5 kg, 0.9 kg, and 0.4 kg, respectively^44^. In our geospatial analysis, higher estimated availability of both fat and carbohydrate was associated with overweight status, alongside higher estimated caloric exposure.

Singapore’s public housing landscape is highly structured with over 80% of residents residing in densely aggregated vertical housing, prioritising short travel distances, extensive public transport connectivity and co-location of residential blocks with food courts, supermarkets and retail clusters. Whilst this enhances accessibility, we did not find any significant correlations between walkability with overweightness, suggesting that associations with BMI are negligible or highly complex interactions are in place. In some settings, higher density of intersections (*n*/km^2^) have been associated with increased walkability and proposed health benefits^45^, yet in others studies contrary findings have been reported. In a study of 233 cities in South America, slightly higher BMI (0.1 units) and increased odds of obesity (4–5%) were observed with increased intersection density although findings were then attenuated with the addition of other built environment features^46^. The same study also found that obesity was negatively associated with obesity. The lack of association of overweightness in Singapore and negative association with green space density could reflect a similar urban structural finding, that more fragmented and greener areas within urban spaces allows for increased physical activity and reduces the influence of the food environment on BMI.

A substantial body of literature has examined the relationship between green space exposure and obesity-related outcomes, generally suggesting protective associations. Many studies measure greenness using satellite-derived indicators such as the Normalized Difference Vegetation Index (NDVI) or by assessing proximity and accessibility to parks and vegetated areas^47^. Longitudinal analyses in the United States^48^ and Europe^49^ have found that higher residential greenness is associated with lower BMI and reduced weight gain over time, while cohort studies in China report lower odds of overweight and central obesity among populations living in greener environments^50^. Similar findings have been observed in large observational studies, including the Nurses’ Health Study, where greater surrounding greenness was linked to higher levels of physical activity and improved health outcomes^51^. Meta-analyses further suggest that access to green space may be associated with approximately 10–15% lower odds of overweight or obesity^52,53^. However, the evidence remains heterogeneous, with some studies reporting null or inconsistent findings depending on how green space is defined and measured. Differences in exposure metrics, including NDVI, park accessibility, or street-level greenery, as well as variations in urban context and population characteristics, likely contribute to these mixed results^52,54,55^. For Singapore, we observed a protective effect with the inclusion of the effects of the food environment. Residents in greener areas in highly urbanised settings are likely to report more walking and recreational activity, and have better cardiometabolic profiles, which could partially offset the effects of high saturated fat intake^53^.

With globalisation, diets have increasingly shifted toward patterns high in both fat and carbohydrate, accompanied by greater availability of energy-dense and nutrient-poor foods^56^. In addition, an expert panel evaluation of Singapore’s food environment highlighted limited implementation of robust local policies and zoning laws targeting unhealthy food environments, alongside limited income support mechanisms that promote access to healthier foods. In this context, heavily discounted quick service and convenience-oriented food options may have minimal incentive to prioritise nutritional quality, instead optimising for palatability and rapid production. Consistent with these observations, our data indicate that areas characterised by higher saturated fat exposures were positively correlated (Table S3), suggesting spatial clustering of energy dense food environments. This pattern may reflect a broader trend across rapidly urbanising Southeast Asian cities, warranting further investigation. For example, Thailand has reported intakes exceeding Planetary Health Diet recommendations for sugar by 452% and saturated oils by 20%, while vegetable intake fell 63% below the recommended lower threshold^57^. In Java, Indonesia, 81.1% of outdoor food and beverage advertisements were found to be advertising unhealthy foods, high in sugars or saturated fats^58^. In Malaysia, a study of 1,173 adults found that 21.3% were obese, yet obesity was not associated with frequenting fast food restaurants^59^. Together, these findings also support the hypothesis that food outlet classification alone may not adequately capture nutritional risk in Southeast Asian settings.

This high-resolution geospatial analysis complements existing dietary intake studies, which are often constrained by limited sampling scale and may be affected by self-reporting bias^60^. It also complements meal-level nutritional assessments, which frequently rely on relatively small samples with limited spatial coverage, and cohort studies that lack sufficiently granular dietary intake measures. In addition, randomised controlled trials may be constrained by sample size and duration, limiting inference on long-term dietary effects. In response to these limitations, new large-scale geospatial and mobility-based approaches are increasingly being explored. These include analyses of 62 million individuals’ visits to food outlets in the United States, where a 10% increase in exposure to fast food outlets in mobile environments increased the odds of visitation by 20%^61^, as well as the development of retail food activity indices using mobility data from 94 million individuals linked to 359,000 food retailers, and diet tracking approaches covering 1.16 million participants^62^. However, most high-resolution geospatial evidence remains concentrated in Western settings such as the United States, the United Kingdom, Australia, and Sweden, with comparatively limited evidence from Southeast Asia^63^.

We propose that food environment metrics should move beyond classifying food outlet types and instead focus on the nutritional composition of foods available within those outlets. While our study was unable to collect food samples from all establishments, the increasing availability of nutritional information online enables more granular characterisation of food environments based on the nutritional content of products offered. This is supported by reviews that has found 70-78% of studies explored found no relationship between the food environment and adult obesity^64^. There is substantial heterogeneity and inconsistency in how food environments are being defined, ranging from fast food classification^64^ to composite food outlets measures combining both healthy and unhealthy food outlets^65^. The application of food environment metrics across multiple settings, especially in understudied locations, is therefore very challenging, limiting comparability across studies and contexts. Future work could incorporate large-scale sampling of food outlets to examine variation in nutritional composition across establishments, discrepancies between reported menu information and actual offerings, and heterogeneity in standard meals across locations. Integrating such data with dietary intake information from population cohorts would allow for more precise assessment of nutritional exposure and its relationship with obesity outcomes.

There are several limitations in our study. Our analysis aims to assess the impact of large-scale patterns of food environment exposures on overweightness and does not capture individual food choices, snacking behaviours or specific diets being carried out at home. We also assume that the majority of meals are consumed within an individual’s neighbourhood food environment. Within food outlets, heterogeneity exists although many standard meals are likely to be very similar in caloric profile. Large-scale sampling of food outlets over an expansive area is unlikely to be feasible, and our method serves as a proxy. Our measure of overweightness using BMI allows for comparison with other studies but further investigations are warranted for hip-waist and hip-height ratios, which are additional measures of obesity. BMI does however serve as an indicator of overconsumption of calories within a large majority of individuals, especially those who are physically inactive, representing a large proportion of the population^66^. Whilst we controlled for multiple confounders, there may be other external influences such as advertising, promotions and popularity of specific outlets in districts that cannot be accounted for without food outlet purchase data. Our macronutrient-based classification into quantiles may also neglect some heterogeneities within zones although it should be noted that the 954 zones match residential clusters and are representative of their local food environments. Our study also did not explore the impact of grocery stores and food purchase decisions within, instead focusing on the eating-out culture which is a common daily practice. The impact of changing food environments also warrants investigation with longer cohorts and the collection of food establishment information over time.

In summary, our findings suggest that higher exposure to energy-dense local food environments, and not the density of fast food outlets, is associated with increased likelihood of overweight status, in Singapore. These results support the need for stronger policy attention to nutritional quality within urban food environments in comparable cities in the region experiencing rising overweight and obesity burdens. Importantly, our findings also emphasise the value of evaluating food environments using macronutrient-based composition and energy content rather than relying solely on outlet classification, since conventional Western definitions of unhealthy food environments may not translate directly to Southeast Asian contexts.

## Data Availability

All data produced in the present study are available upon reasonable request to the authors

## Notes

### Competing Interest Statement

The authors have declared no competing interest.

### Funding Statement

This study was funded by NUS Ground Up Seed Fund (A-8001108-00-00).

### Author Declarations

The Institutional Review Board of the National University of Singapore gave ethical approval for this work (NUS-IRB-2022-794).

### Summary of Updates

Figures revised; author affiliations updated; Supplemental files updated.

