## Supplementary Information for "Planning healthier cities by reframing the urban food landscape: measuring localised macronutrient exposure in Singapore"

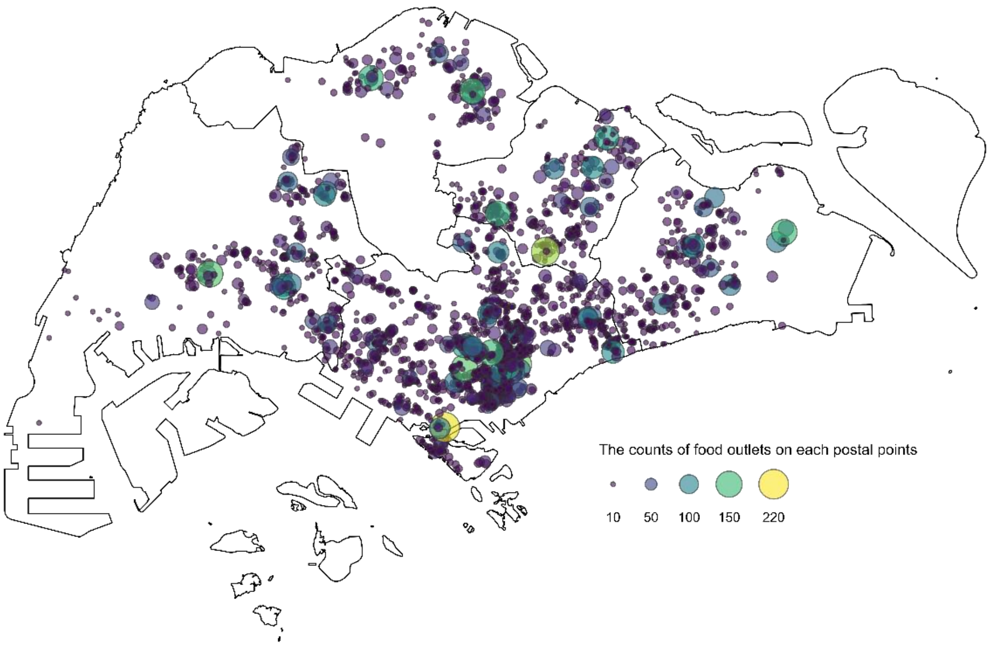

**Supplementary Figure S1 | Spatial distribution of 14,764 geolocated food establishments across Singapore.** Individual postal codes are represented by circles, with larger circles and lighter colours indicating a higher number of food establishments co-located at the same address.

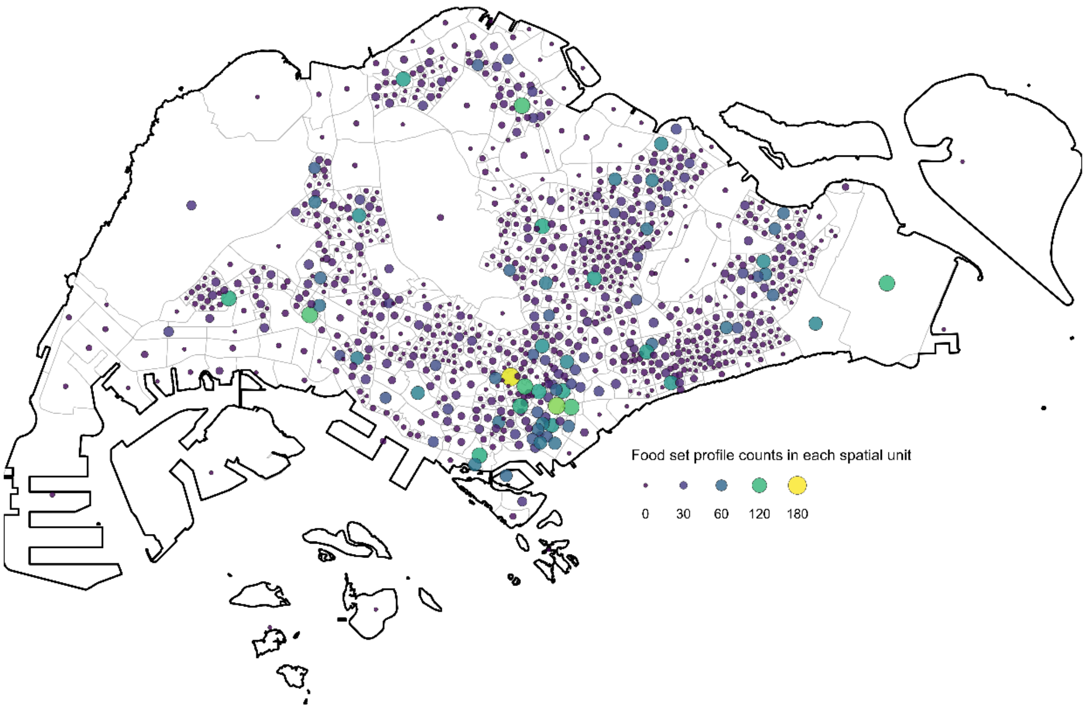

**Supplementary Figure S2 | Spatial distribution of food set profile counts across Singapore.** Circles located at the centroid of each spatial unit represent the number of distinct food set profiles available, with larger sizes and lighter colours indicating higher counts.

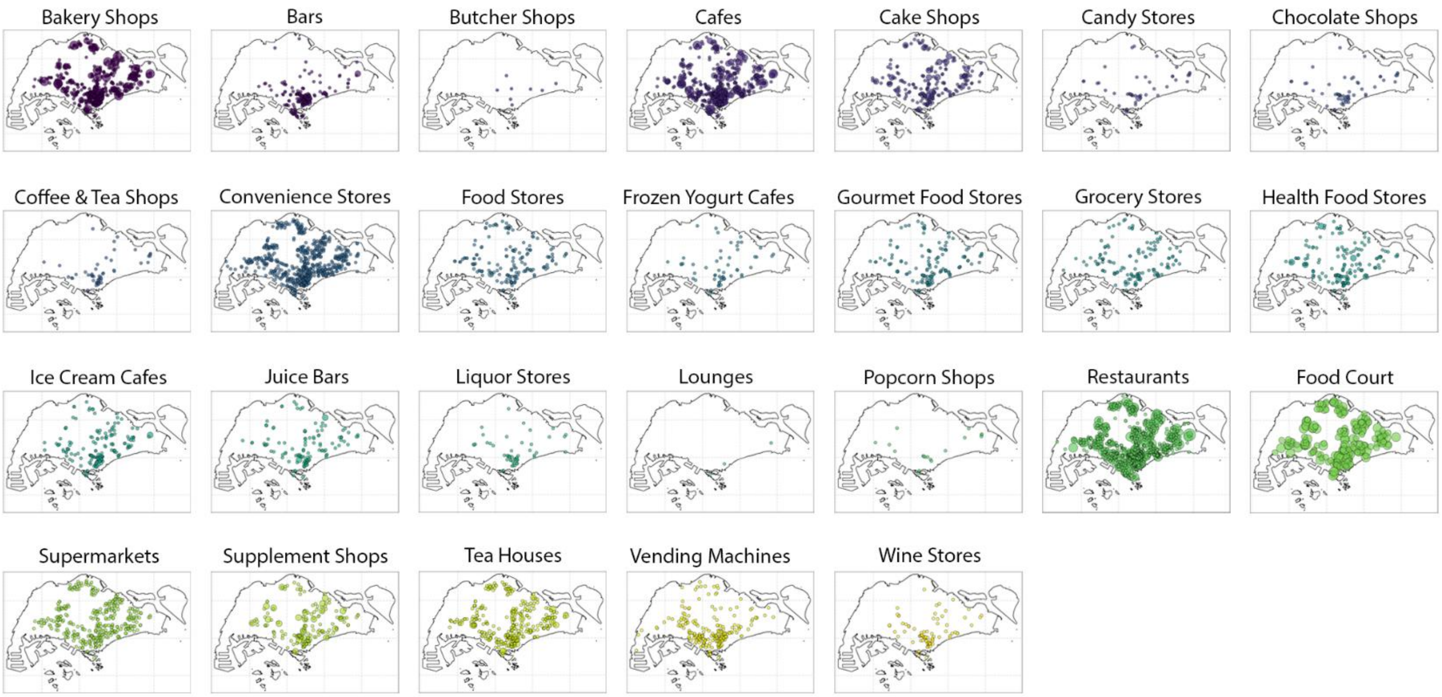

**Supplementary Figure S3 | Spatial distribution of food establishments by retailer type in Singapore.** In each panel, circles represent individual postal codes, with larger circles and lighter colours indicating a higher number of food establishments co-located at the same address within each retailer type.

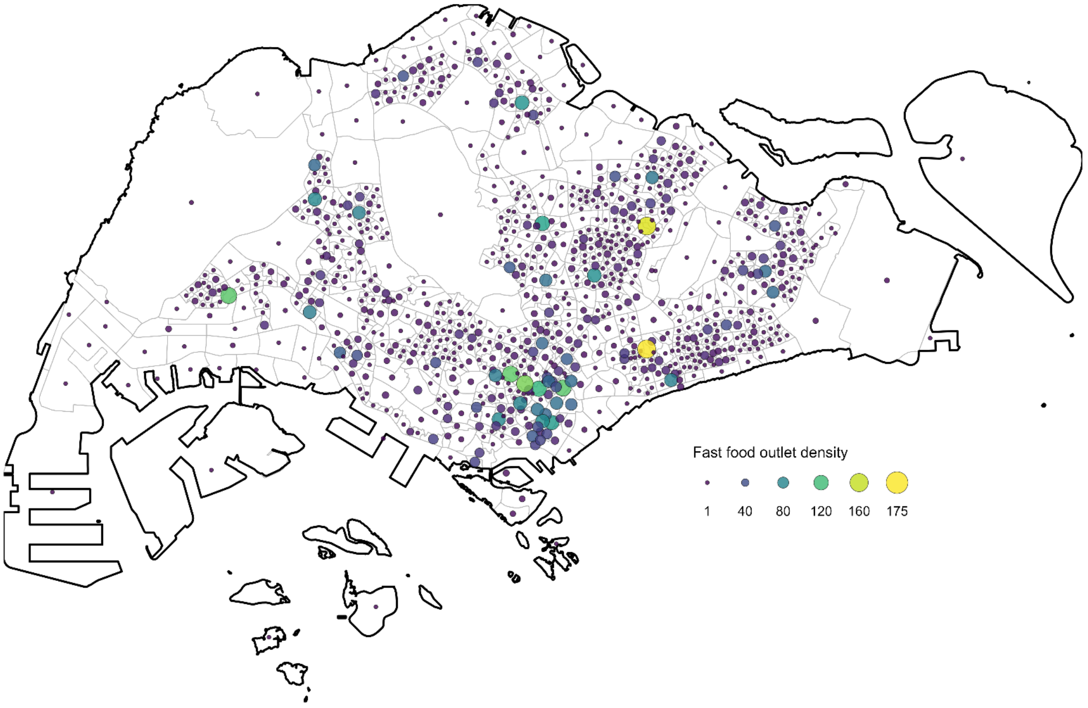

**Supplementary Figure S4 | Spatial distribution of fast food outlets density in Singapore.** Circles located at the centroid of each spatial unit present the density of fast food outlets per square meter, with larger sizes and lighter colours indicating higher density.

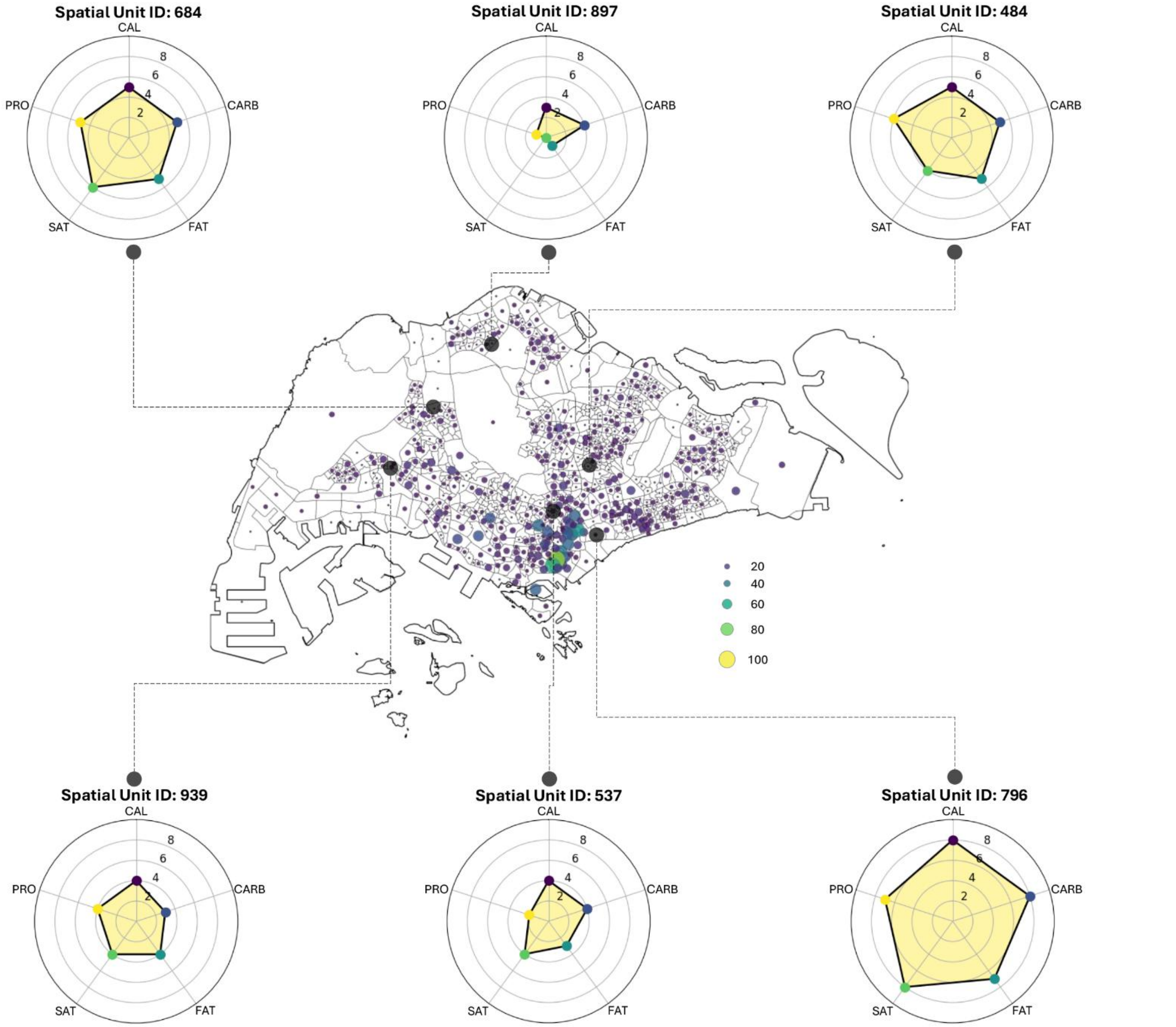

**Supplementary Figure S5 | Nutrient exposure across spatial units in Singapore.** The central map presents the spatial distribution of the number of postal codes containing food establishments within each spatial unit across Singapore, where larger circles and lighter colours indicate higher establishment density. The six radar plots visualise the values of nutrient-related indices (integer-valued, ranging from 0 to 10) for selected spatial units across the city.

**Supplementary Table S1 | Summary of socio-demographic and anthropometric characteristics of the 15,614 participants, stratified by overweight status.**

| | Total (N = 15,614) | Overweight (N = 9548) | Non-overweight (N = 6066) | P-value^ |
|---|---|---|---|---|
| **Demographics** | | | | |
| **Age (years)** | | | | |
| 21-24 | 264(1.7%) | 110(1.2%) | 154(2.5%) | <0.001 |
| 25-34 | 2149(13.8%) | 1101(11.5%) | 1048(17.3%) | |
| 35-44 | 3282(21.0%) | 2060(21.6%) | 1222(20.1%) | |
| 45-54 | 4235(27.1%) | 2780(29.1%) | 1455(24.0%) | |
| 55-64 | 3503(22.4%) | 2214(23.2%) | 1289(21.2%) | |
| 65+ | 2181(14.0%) | 1283(13.4%) | 898(14.8%) | |
| **Gender** | | | | |
| Male | 7151(45.8%) | 4806(50.3%) | 2345(38.7%) | <0.001 |
| Female | 8463(54.2%) | 4742(49.7%) | 3721(61.3%) | |
| **Ethnicity** | | | | |
| Chinese | 10429(66.8%) | 5526(57.9%) | 4903(80.9%) | <0.001 |
| Malays | 2365(15.1%) | 1810(18.9%) | 555(9.1%) | |
| Indians | 2820(18.1%) | 2212(23.2%) | 608(10.0%) | |
| **Marital status** | | | | |
| Single | 2960(19.0%) | 1445(15.2%) | 1515(25.0%) | <0.001 |
| Married | 11201(71.7%) | 7183(75.2%) | 4018(66.2%) | |
| Divorced | 72(0.5%) | 41(0.4%) | 31(0.5%) | |
| Separated | 898(5.7%) | 542(5.7%) | 356(5.9%) | |
| Widowed | 483(3.1%) | 337(3.5%) | 146(2.4%) | |
| **Social-economic Status** | | | | |
| **Monthly household income (SGD)** | | | | |
| 0 to <2000 | 3976(25.5%) | 2429(25.5%) | 1547(25.5%) | <0.001 |
| 2000 to <4000 | 4368(27.9%) | 2754(28.9%) | 1614(26.6%) | |
| 4000 to <6000 | 3131(20.1%) | 1961(20.5%) | 1170(19.3%) | |
| 6000+ | 2703(17.3%) | 1560(16.3%) | 1143(18.8%) | |
| Refused/Don't know/Missing | 1436(9.2%) | 844(8.8%) | 592(9.8%) | |
| **Education level** | | | | |
| A-level, equivalent, or lower | 12040(77.1%) | 7538(78.9%) | 4502(74.2%) | <0.001 |
| University or above | 3574(22.9%) | 2010(21.1%) | 1564(25.8%) | |
| **Education duration (years)** | | | | |
| 0–10 | 6242(40.0%) | 4004(41.9%) | 2238(36.9%) | <0.001 |
| 11+ | 9372(60.0%) | 5544(58.1%) | 3828(63.1%) | |
| **Work status** | | | | |
| Working | 11407(73.1%) | 7067(74.0%) | 4340(71.5%) | <0.001 |
| Student | 422(2.7%) | 162(1.7%) | 260(4.3%) | |
| Homemaker | 2301(14.7%) | 1474(15.4%) | 827(13.6%) | |
| Retired | 834(5.3%) | 473(5.0%) | 361(6.0%) | |
| Unemployed | 650(4.2%) | 372(3.9%) | 278(4.6%) | |
| **Housing type** | | | | |
| HDB# 1–2 room flat | 1413(9.0%) | 906(9.5%) | 507(8.4%) | 0.28 |
| HDB# 3 room flat | 3537(22.7%) | 2129(22.3%) | 1408(23.2%) | |
| HDB# 4 room flat | 6028(38.6%) | 3691(38.7%) | 2337(38.5%) | |
| HDB# 5 room or executive flat | 4445(28.5%) | 2704(28.3%) | 1741(28.7%) | |
| Private property | 191(1.2%) | 118(1.2%) | 73(1.2%) | |
| **Physical Health** | | | | |
| **Smoking Status** | | | | |
| Never smoked | 11933(76.4%) | 7100(74.4%) | 4833(79.7%) | <0.001 |
| Ex-smoker | 1224(7.9%) | 877(9.2%) | 347(5.7%) | |
| Current occasional smoker | 457(2.9%) | 305(3.2%) | 152(2.5%) | |
| Current daily smoker | 2000(12.8%) | 1266(13.2%) | 734(12.1%) | |
| **Hypertension** | | | | |
| Yes | 2668(17.1%) | 2075(21.7%) | 593(9.8%) | <0.001 |
| No | 12946(82.9%) | 7473(78.3%) | 5473(90.2%) | |
| **Total physical activity level** (met-hr/wk) | | | | |
| 0 to <50 | 2905(18.6%) | 1615(16.9%) | 1290(21.3%) | <0.001 |
| 50 to <100 | 5134(32.9%) | 3055(32.0%) | 2079(34.3%) | |
| 100 to <150 | 3733(23.9%) | 2362(24.7%) | 1371(22.6%) | |

| | | | |
|---|---|---|---|
| 150+ | 3842(24.6%) | 2516(26.4%) | 1326(21.8%) |

^ Between-group comparisons were performed using chi-square tests.

[#] Public housing types in Singapore

**Supplementary Table S2| Estimated adjusted odds ratios (posterior medians and 95% CrIs) for socio-demographic predictors from the baseline model and nutrient-adjusted Models 1–6.** In addition to the listed predictors and abundance of green space, Models 1–6 were also adjusted for the calorie index, carbohydrate index, fat index, protein index, saturated fat index, and high saturated fat score, respectively.

| Predictor | Baseline | Model1 | Model2 | Model3 | Model4 | Model5 | Model6 |
|---|---|---|---|---|---|---|---|
| **Gender** | | | | | | | |
| Male (baseline) | 1 | 1 | 1 | 1 | 1 | 1 | 1 |
| Female | 0.57 [0.52,0.61] | 0.57 [0.53,0.61] | 0.57 [0.52,0.61] | 0.57 [0.53,0.61] | 0.57 [0.52,0.61] | 0.57 [0.53,0.61] | 0.56 [0.52,0.61] |
| **Age** | | | | | | | |
| 21–24 | 0.64 [0.47,0.87] | 0.63 [0.46,0.86] | 0.63 [0.46,0.86] | 0.63 [0.46,0.85] | 0.63 [0.46,0.86] | 0.63 [0.46,0.86] | 0.64 [0.47,0.86] |
| 25–34 | 0.87 [0.74,1.01] | 0.86 [0.73,1.00] | 0.86 [0.73,1.01] | 0.86 [0.73,1.01] | 0.86 [0.73,1.01] | 0.86 [0.73,1.00] | 0.86 [0.73,1.01] |
| 35–44 | 1.45 [1.25,1.67] | 1.44 [1.25,1.66] | 1.44 [1.25,1.67] | 1.44 [1.24,1.67] | 1.44 [1.25,1.67] | 1.44 [1.25,1.66] | 1.44 [1.25,1.67] |
| 45–54 | 1.55 [1.35,1.76] | 1.53 [1.35,1.75] | 1.54 [1.35,1.75] | 1.54 [1.34,1.75] | 1.54 [1.34,1.76] | 1.54 [1.35,1.75] | 1.54 [1.34,1.75] |
| 55-64 | 1.33 [1.17,1.51] | 1.33 [1.17,1.51] | 1.33 [1.17,1.51] | 1.33 [1.17,1.51] | 1.33 [1.17,1.51] | 1.33 [1.17,1.51] | 1.33 [1.17,1.51] |
| 65+ (baseline) | 1 | 1 | 1 | 1 | 1 | 1 | 1 |
| **Ethnicity** | | | | | | | |
| Chinese (baseline) | 1 | 1 | 1 | 1 | 1 | 1 | 1 |
| Malays | 3.01 [2.70,3.37] | 3.01 [2.69,3.36] | 3.01 [2.69,3.37] | 3.01 [2.70,3.37] | 3.01 [2.69,3.37] | 3.01 [2.69,3.37] | 3.01 [2.69,3.37] |
| Indians | 3.42 [3.09,3.78] | 3.39 [3.06,3.75] | 3.39 [3.06,3.76] | 3.39 [3.06,3.76] | 3.39 [3.06,3.76] | 3.39 [3.06,3.75] | 3.38 [3.06,3.76] |
| **High education** | | | | | | | |
| No (baseline) | 1 | 1 | 1 | 1 | 1 | 1 | 1 |
| Yes | 0.88 [0.81,0.95] | 0.88 [0.81,0.96] | 0.88 [0.82,0.96] | 0.88 [0.82,0.96] | 0.88 [0.82,0.96] | 0.88 [0.81,0.96] | 0.88 [0.81,0.96] |
| **Work status** | | | | | | | |
| Working (baseline) | 1 | 1 | 1 | 1 | 1 | 1 | 1 |
| Student | 0.67 [0.53,0.85] | 0.67 [0.53,0.85] | 0.67 [0.53,0.85] | 0.67 [0.53,0.85] | 0.67 [0.53,0.85] | 0.67 [0.53,0.85] | 0.67 [0.53,0.84] |
| Homemaker | 1.17 [1.05,1.30] | 1.17 [1.05,1.30] | 1.17 [1.05,1.30] | 1.17 [1.05,1.30] | 1.17 [1.05,1.30] | 1.17 [1.05,1.30] | 1.17 [1.05,1.30] |
| Retired | 0.80 [0.67,0.95] | 0.80 [0.68,0.96] | 0.81 [0.68,0.96] | 0.81 [0.68,0.95] | 0.80 [0.68,0.96] | 0.80 [0.68,0.95] | 0.80 [0.67,0.95] |
| Unemployed | 0.76 [0.64,0.91] | 0.76 [0.64,0.91] | 0.76 [0.64,0.91] | 0.76 [0.64,0.91] | 0.76 [0.64,0.90] | 0.76 [0.64,0.91] | 0.76 [0.64,0.91] |
| **Hypertension** | | | | | | | |
| No (baseline) | 1 | 1 | 1 | 1 | 1 | 1 | 1 |
| Yes | 2.73 [2.45,3.05] | 2.73 [2.46,3.05] | 2.73 [2.45,3.05] | 2.73 [2.45,3.04] | 2.73 [2.45,3.05] | 2.73 [2.46,3.05] | 2.74 [2.45,3.05] |

**Supplementary Table S3 | Estimated adjusted odds ratios (posterior medians and 95% CrIs) for the food and built environment predictors in Model 1–6 further adjusted for clustering effects.** These models respectively incorporated the calorie index, carbohydrate index, fat index, protein index, saturated fat index, and high saturated fat score as predictors in addition to socio-demographic factors and abundance of green space, while also accounting for cluster heterogeneity and spatial autocorrelation.

| Predictor | Model 1 | Model 2 | Model 3 | Model 4 | Model 5 | Model 6 |
|---|---|---|---|---|---|---|
| High green space coverage | 0.71 [0.59,0.77] | 0.76 [0.65,0.85] | 0.72 [0.62,0.79] | 0.78 [0.71,0.83] | 0.73 [0.63,0.80] | 0.48 [0.35,0.57] |
| Calorie index | 1.01 [0.96,1.07] | | | | | |
| Carbohydrate index | | 1.00 [0.98,1.04] | | | | |
| Fat index | | | 1.03 [0.99,1.10] | | | |
| Protein index | | | | 1.02 [0.97,1.07] | | |
| Saturated fat index | | | | | 1.01 [0.96,1.07] | |
| High saturated fat score | | | | | | 1.55 [1.29, 2.49] |
